# Well-being biomarkers and psychological functioning of adult patients during chemotherapy treatment: the effects of hospital clowns and hosting conditions

**DOI:** 10.1101/2023.11.22.23294770

**Authors:** Marta Simões, Albertina Oliveira, Rosário Pinheiro, Manuela Vilar, Diana Agante, Isabel Pazos, Bárbara Oliveiros, Manuela Grazina

**Affiliations:** CIBB – Center for Innovative Biomedicine and Biotechnology (www.cibb.uc.pt/), Coimbra, Portugal; Laboratory of Mitochondrial Biomedicine and Theranostics, CNC – Center for Neuroscience and Cell Biology, University of Coimbra, Coimbra, Portugal; Faculty of Psychology and Education Sciences, University of Coimbra, Coimbra, Portugal; CEIS20 Centre for Interdisciplinary Studies, University of Coimbra, Portugal; CINEICC The Center for Research in Neuropsychology and Cognitive and Behavioral Intervention, Coimbra, Portugal; IPO – Portuguese Institute of Oncology of Coimbra, Coimbra, Portugal; FMUC – Faculty of Medicine, University of Coimbra, Coimbra, Portugal

**Keywords:** Well-being, biomarkers, cortisol, psychological functioning, ambulatory cancer chemotherapy, hospital clowns

## Abstract

**Background:** Pharmacological oncological treatments interfere with the patient’s quality of life on physical, psychological, and social dimensions. Besides all the care in hosting conditions, hospital clowns (HCs) use artistic sketches aiming to reduce distress, but studies analyzing their effects on biomarkers in association with psychological states are scarce. This study examined biomarkers and psychological functioning related to its effects, in adult patients of an ambulatory chemotherapy hospital setting.

**Methods:** Following a pilot study with pre-testing/post-testing, 64 women were analysed: experimental group (EG; n=36; HCs intervention plus hosting conditions) *versus* control group (CG; n=28; hosting conditions). Oxytocin, cortisol, serotonin and ATP saliva levels were measured. Psychological assessment covered positive and negative affect (PA; NA), emotional states (PESS), mindfulness attention’ quality (CAMS-R), cognitive functioning (CDT) and satisfaction with HCs.

**Results:** Data did not reveal statistically significant differences in biomarkers between groups; EG showed improvements in psychological functioning, in which NA had significantly decreased, compared to CG; PESS and CAMS-R have also improved. Association analyses suggest a role of serotonin in moderating the relationship between (PA&scholarity)&CAMS-R with NA&age; serotonin and ATP changes are more related to psychological features; PESS seems to modulate those relationships in EG.

**Conclusions:** Although similar results were found for the two groups in study, concerning biomarkers, the psychological functioning showed that short-term interventions during ambulatory chemotherapy might increase well-being of adult cancer patients. Certainly, a larger sample is needed, both to ascertain salivary biomarkers variations and psychological benefits, but this study is undoubtedly pioneer.

## Introduction

Well-being is characterized by feelings of trust, strong social networks and ability to relate with others. Nevertheless, fear, anxiety, isolation, lack of social support as well as physiological risk factors, such as high blood pressure and release of stress hormones, are harmful factors to emotional and physical well-being (1, 2). Oncological diseases are one of the most disruptive and challenging life events, triggering high negative symptoms. In addition to the shock of the diagnosis, physical symptoms (e.g., pain, fatigue, nausea, etc.) and high psychological stress affect the dynamic balances of well-being and negatively influence the recovery process and quality of life of oncological patients (3, 4).

Among the theories explaining the perception of well-being in adulthood, the conceptualizations around subjective well-being by Diener (5) stands out. This model includes a cognitive dimension related to cognitive judgments about life satisfaction and an affective component regarding emotional reactions to life events, the latter divided into positive and negative affect (6). In general, people with high positive affectivity deal better with stress-inducing situations and perceive themselves to be more in control of their own lives. Within the scope of this theory, strategies that increase positive affect and decrease negative affect improve the perception of well-being.

Besides the comfort given by hospital’s hosting conditions, arts’ activities can be considered as complex interventions that combine different components, such as the involvement of the imagination, sensory activation, evocation of emotion and cognitive stimulation. In addition, they also involve social interaction, physical activity, engagement and interaction with health issues/environments. Social interaction can reduce loneliness and lack of social support, which are both linked with adverse physiological responses, cognitive, functional and motor decline (7). Studies focusing on humor and laughter (e.g., 8) have shown that these could be assumed as a coping strategy to deal with stress (9–13). In the hospital context, particularly in the case of highly stressful illnesses, like cancer, humor and laughter have gained importance as tools to reduce the negative impact of the illness. This approach enhances positive emotions, allowing patients to deal better with their condition (10, 14), although the majority of studies comes from pediatric settings (e.g., 15-17). The main purpose of Hospital Clowns (HCs) is to promote the well-being of all individuals with whom they interact. The HCs are artistic performers trained to generate a positive atmosphere at the Hospital (18), developing warm interpersonal relations with patients and health staff through play and laughter (13, 19). Stimulating interaction through creative performance, they are also trained to capture patients’ attention to the present moment, helping to focus and being aware of what’s happening in the moment and thus distracting them from the negative impact of disease stressors (19–21). However, quantitative studies appraising its impact in human health and well-being are still missing.

Research in the fields of behavioral and psychosomatic medicine revealed associations of psychological well-being levels with subsequent physical health outcomes (e.g., 22). Accordingly, the analysis of several biomarkers can be correlated to well-being, in order to evaluate the impact of HCs intervention, besides to hosting conditions, in biochemical parameters.

It is well known that oxytocin is synthesized in hypothalamus with projections to neurohypophysis and sites within the central nervous system (23), playing a key role in fear conditioning, social cognition, social behaviors and affiliation and maternal bonding (24). It has also an important role in social anxiety and impaired social functioning disorders; in clinical reports, oxytocin was related to psychiatric disorders like depression, autism, anxiety disorder and post-traumatic stress disorder (25–28).

Cortisol is an adrenal hormone released in response to stress (29), being a well-established biomarker of psychological stress and related mental or physical diseases (30–33). A response to chronic stress with prolonged cortisol secretion may increase individual’s susceptibility to poor health and disease (34–37).

Serotonin is a neurotransmitter synthesized in brain, platelets, intestinal myenteric plexus and gut enterochromaffin cells (38, 39). It is well known for its role in mood regulation and impact in numerous peripheral physiological functions (40). Several studies have shown an association between a trait of empathy and helping behavior, such like care for the baby’s crying (41). Serotonin system may be involved in individual empathic abilities, since it plays a key role in modulating emotional states (42).

Furthermore, ATP biology is intrinsic to mitochondria, which own signaling enabling stress adaptation (43). An emerging concept proposes that mitochondria detect, integrate and translate psychosocial and behavioral factors into cellular and molecular changes, and in turn contribute to the biological embedding of psychological states. Clinical features of mitochondrial defects indicate that both the hypothalamic-pituitary-adrenal (HPA) and sympathetic-adrenal-medullary (SAM) axes activities may be directly modulated by mitochondrial function (43, 44). Chronic stress-induced mitochondrial allostatic load lowers mitochondrial bioenergetics capability and increases depressive mood states, as depressive symptoms can ameliorate by enhancing mitochondrial function, including ATP synthesis (45).

Accordingly, the detection of quoted substances in biological fluids have gained importance in emotional state indicators analysis (46, 47), being potential biomarkers of well-being.

Therefore, an innovative approach is herein presented, concerning the implementation, evaluation and monitoring methodology, in order to assess biomarkers of the well-being status. To our knowledge, this is the first study assessing changes in biomarkers and psychological symptoms in adults with cancer, in particular related to clown performances in treatment rooms.

## Methods and Materials

### Study design

A pilot study with a factorial repeated measures design (group x time) was implemented, in order to evaluate biomarkers’ levels and psychological functioning upon HCs intervention. “Group” refers to participants’ exposure (intervention plus hosting conditions – experimental group, EG, and hosting conditions only – control group, CG); “time” refers to repeated measures (pre– and post-test). Pre-test and post-test procedure was similar for all participants, saliva collection followed by psychological assessment, considering the intervention’s duration (see Supplemental Information).

### Participants, Recruitment, and Procedures

This study was carried out at ambulatory service (See Supplemental Information) in the Portuguese Institute of Oncology of Coimbra (IPO). A sample of 64 subjects was included in the study (n=36, EG; n=28, CG), evaluated in two moments (basal and after 90 minutes (intervention for EG). In order to avoid gender bias related to variations in the parameters in study, only adult women were included, aged 57±12 (32–90) years old.

Data were obtained after participants signing informed consent, according to Law no.12/2005, DL no.131/2014 and the Norm 015/2013 of the Directorate-General of Health (DGS), following the Tenets of the Helsinki Declaration. Samples were anonymized following the European Union General Data Protection Regulation (GDPR – EU 2016/679) and the Portuguese Law (Law no.58/2019).

Supplementary table (S1) contains sociodemographic data, including age, educational level, marital status and profession.

Detailed information on samples’ collection, determination of biomarkers’ levels and psychological evaluation is detailed in Supplemental Information.

### Intervention

The HCs “Palhaços D’Opital (PdO)” have a program aiming to bring joy, affection and good mood to adults and elders in hospital environment. They work in pairs, dressed in colorful costumes and they prepared sketches in advance. Their performances recreate day-to-day situations (e.g., clowns trying to install a new TV screen; planning a wedding between the clowns and asking the public for their ideas), ending with specific closure, in order to indicate to the audience that the session had finished and to promote a positive environment after their departure. Each intervention lasted around 15 minutes.

## Results

### Effect on biomarkers

The variation in biomarkers concentration before and after the HCs’ intervention – EG – (or equivalent waiting time – CG) was analyzed, in order to assess the impact at the biochemical level. In the case of oxytocin, serotonin and ATP, there was no pre-post evaluation variation in concentration (Table 1). A statistically significant effect was found for cortisol levels, in pre-post evaluation (p=0.045) in the overall sample (both groups), with an estimated overall mean decrease of 1.0 ± 0.5 units, although the lack of statistical significance in the comparison between EG and CG, paired for time of observation (Table 1), with higher difference observed in the CG.

**Table 1:**
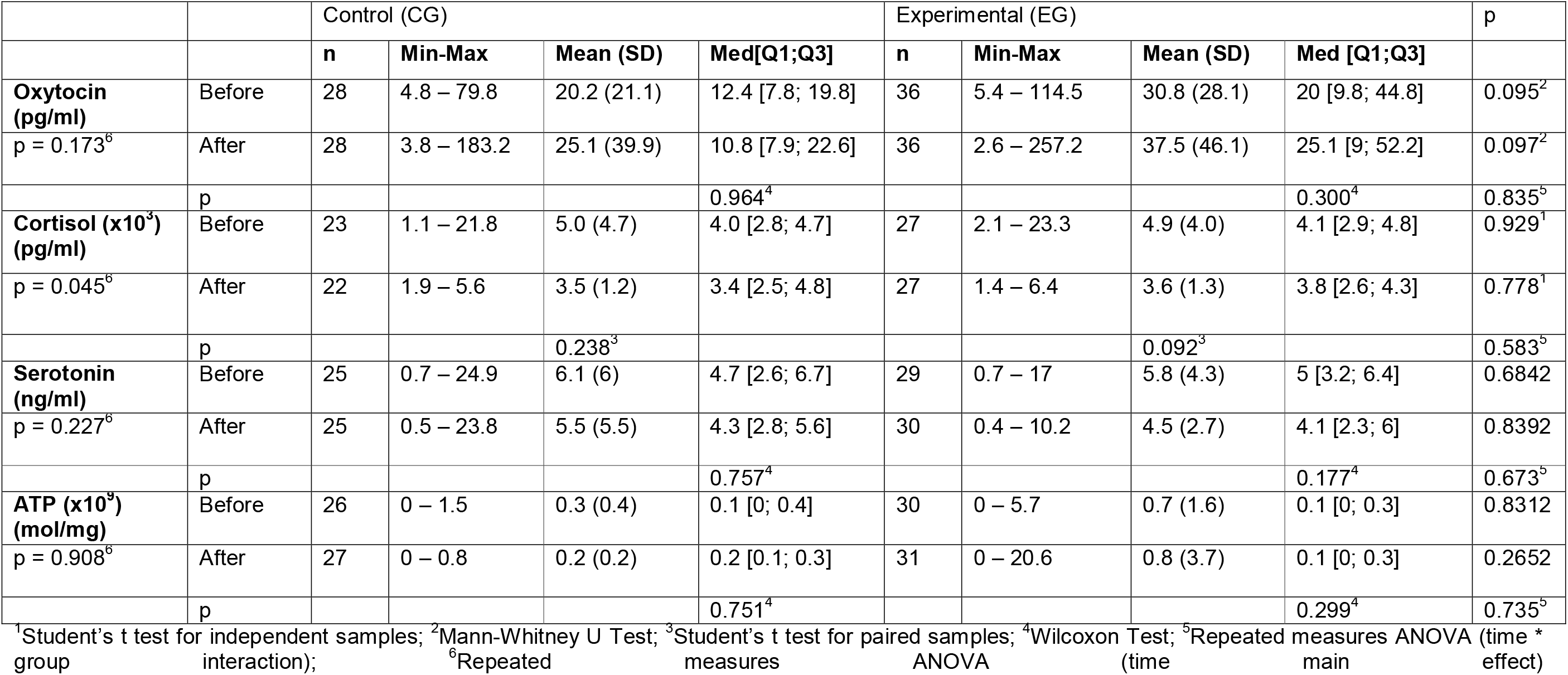
Descriptive statistics of biomarkers evaluation and comparisons between groups.

The results reveal no statistically significant differences concerning the effect of time, group or interaction concerning biomarkers.

### Effect on psychological evaluation

Concerning interaction between group and moment of evaluation, a statistically significant effect on NA was found (p=0.032). There was a statistically significant difference between CG and EG either for pre-test (p=0.042) or post-test evaluation (p<0.001), with a higher value for CG. There was an overall decrease in the values between both evaluations (p=0.013) attributed to the difference presented by EG (p=0.001) [Table 2, Figure 1A]. No statistically significant interaction was found for other psychological characteristics evaluated.

**Figure 1.**
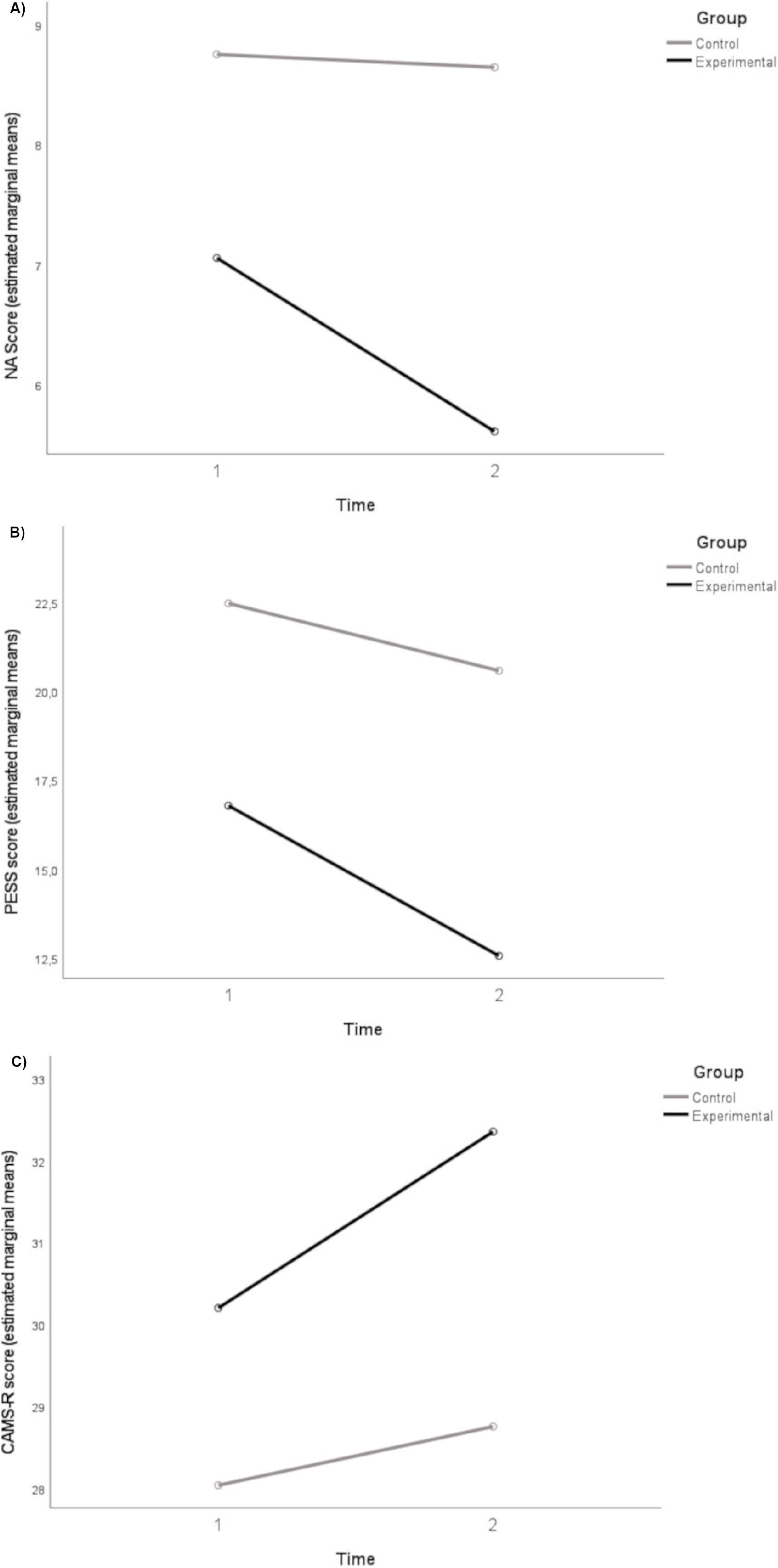
– Psychological functioning variation between pre– and post-test evaluations for both CG and EG. A) NA (no statistically significant variation found in the CG – p = 0.622 – but there is statistical evidence for differences in the EG – p=0.001); B) PESS (statistically significant variation found for the EG – p <0.001. No evidence of statistically significant variation in CG – p=0.068); C) CAMS-R (statistically significant variation found for the EG – p=0.037 – but no evidence of statistically significant variation in the CG – p = 0.585). CAMS-R, Cognitive and Affective Mindfulness measured by the Cognitive and Affective Mindfulness Scale-Revised, NA, Negative Affectivity assessed by the Positive and Negative Affect Schedule (PANAS), PESS, Emotional States assessed by the Perception of Emotional States Scale.

**Table 2:** Descriptive statistics of psychological evaluation and comparisons between groups.

|  |  | Control (CG) |  |  |  | Experimental (EG) |  |  |  | p |
| --- | --- | --- | --- | --- | --- | --- | --- | --- | --- | --- |
|  |  | n | Min-Max | Mean (SD) | Med [Q1; Q3] | n | Min-Max | Mean (SD) | Med [Q1; Q3] |  |
| **CDT**<br>p = 0.432<sup>6</sup> | Pre-test | 26 | 6 – 17 | 13.8 (2.8) | 15 [14; 15] | 32 | 5 – 18 | 13.4 (4) | 15 [9; 17] | 0.664<sup>2</sup> |
|  | Post-test | 26 | 8 - 17 | 14 (2.2) | 14.5 [14; 15] | 27 | 1 – 18 | 14.4 (4.3) | 16 [14; 17] | 0.037<sup>2</sup> |
|  | P |  |  |  | 1.000<sup>4</sup> |  |  |  | 0.442<sup>4</sup> | 0.702<sup>5</sup> |
| **PESS**<br>p < 0.001<sup>6</sup> | Pre-test | 28 | 8 - 47 | 22.5 (10.7) | 23 [14; 26.5] | 36 | 8 – 27 | 16.8 (5.3) | 17.5 [13.5; 19.5] | 0.014<sup>1</sup> |
|  | Post-test | 28 | 8 - 45 | 20.6 (9.7) | 20.5 [13; 26.5] | 36 | 8 – 20 | 12.6 (3.9) | 12 [8.5; 16] | <0.001<sup>1</sup> |
|  | P |  |  | 0.068<sup>3</sup> |  |  |  | < 0.001<sup>3</sup> |  | 0.074<sup>5</sup> |
| **PA**<br>p = 0.996<sup>6</sup> | Pre-test | 28 | 10 - 24 | 16 (3.4) | 16 [13.5; 18] | 36 | 10 – 24 | 16.8 (3.3) | 16 [14.5; 18.5] | 0.345<sup>1</sup> |
|  | Post-test | 28 | 9 - 25 | 15.3 (4.4) | 14.5 [11.5; 19] | 36 | 11 – 25 | 17.4 (3.4) | 17 [15; 20] | 0.036<sup>1</sup> |
|  | P |  |  | 0.211<sup>3</sup> |  |  |  | 0.277<sup>3</sup> |  | 0.111<sup>5</sup> |
| **NA**<br>p = 0.013<sup>6</sup> | Pre-test | 28 | 5 - 19 | 8.8 (3.6) | 8 [5.5; 10.5] | 36 | 5 – 17 | 7.1 (2.6) | 6 [5; 7.5] | 0.042<sup>2</sup> |
|  | Post-test | 28 | 5 - 21 | 8.6 (4.1) | 7.5 [5; 10] | 36 | 5 – 11 | 5.6 (1.2) | 5 [5; 6] | <0.001<sup>2</sup> |
|  | P |  |  |  | 0.622<sup>4</sup> |  |  |  | 0.001<sup>4</sup> | 0.032<sup>5</sup> |
| **CAMS-R**<br>p = 0.044<sup>6</sup> | Pre-test | 28 | 13 - 23 | 16.8 (2.9) | 17 [14.5; 19] | 36 | 12 – 24 | 18.1 (2.6) | 18 [16; 20] | 0.059<sup>1</sup> |
|  | Post-test | 28 | 9 - 22 | 17.1 (2.9) | 17 [15; 19] | 29 | 14 – 27 | 19.5 (3.2) | 19 [17; 21] | 0.005<sup>1</sup> |
|  | P |  |  | 0.585<sup>3</sup> |  |  |  | 0.037<sup>3</sup> |  | 0.173<sup>5</sup> |

<sup>1</sup> Student's t-test for independent samples; <sup>2</sup> Mann-Whitney U-Test; <sup>3</sup> Student's t test for paired samples; <sup>4</sup> Wilcoxon Test; <sup>5</sup> Repeated measures ANOVA (time \* group interaction); <sup>6</sup> Repeated measures ANOVA (time main effect).
CAMS-R, Cognitive and Affective Mindfulness measured by the Cognitive and Affective Mindfulness Scale-Revised, CDT, Cognitive functioning assessed by the Clock-Drawing Test, NA and PA, Affectivity assessed by the Positive and Negative Affect Schedule (PANAS), PESS, Emotional States assessed by the Perception of Emotional States Scale.

Regarding both moments of evaluation, a statistically significant difference was found for PESS (p< 0.001) and CAMS-R (p=0.044), due to differences in the EG (PESS: p< 0.001; CAMS-R: p=0.037) and between groups, particularly in the post-test evaluation: pre and post-test comparisons between groups – PESS: p=0.014 and p<0.001; CAMS-R: p=0.059 and p=0.005 (Table 2). There was an increase in CAMS-R (EG with higher values than CG) and a decrease in PESS between both evaluations (EG presented lower values than CG) (Figure 1 – B, C).

Both CG and EG presented statistically significant differences in PA and CDT in post-test (p=0.036 and p=0.037, respectively; Table 2, Figure S1-D, E).

### Associating biomarkers’ levels and psychological evaluation

Data correlation analysis (Table 3) revealed statistically significant results for cortisol/ATP levels variation in the EG (r=0.357) and between serotonin and both oxytocin or ATP variations in the CG (r=0.336, r=0.456, respectively). Additionally, there were a few moderate correlations for the psychological parameters in the EG (PA correlated negatively to CDT and positively to CAMS-R changes (r=-0.449, r=0.491, respectively), whereas NA was positively correlated to PESS (r=0.523). On the other hand, there were statistically significant negative correlations between oxytocin and both PA and CAMS-R changes in the CG (r=-0.355, r=-0.433, respectively).

**Table 3:**
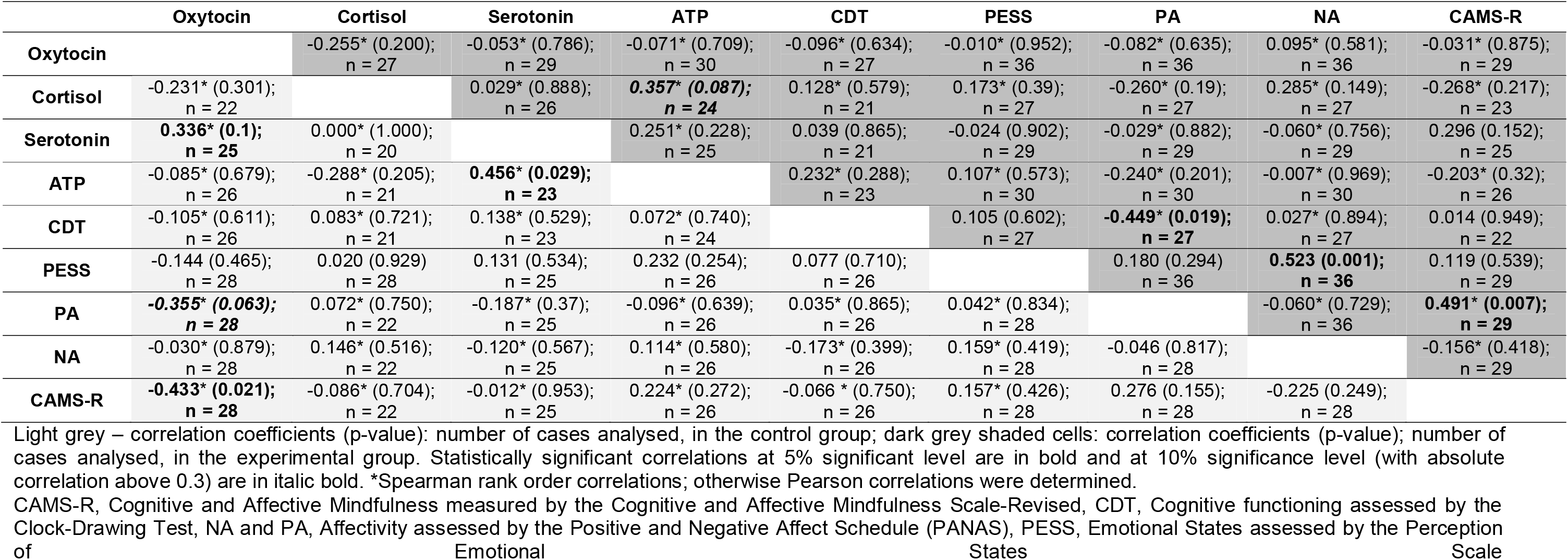
Correlation between biomarkers’ and psychological variables, pre and post-test evaluation.

A hierarchical cluster analysis on variables using the Manhattan distance and single linkage identifies clusters as follows (Figure 2): the variation in PA modulated by the education level (scholarity) and CAMS-R are related, similarly to the impact of age in NA. It also shows that serotonin may play a role in moderating the relationship between the clusters identified above ((PA&scholarity) & CAMS-R with NA&age). Moreover, CDT and PESS are closely related (Figure 2). Separated group analyses can be seen in supplemental Figure S2.

**Figure 2.**
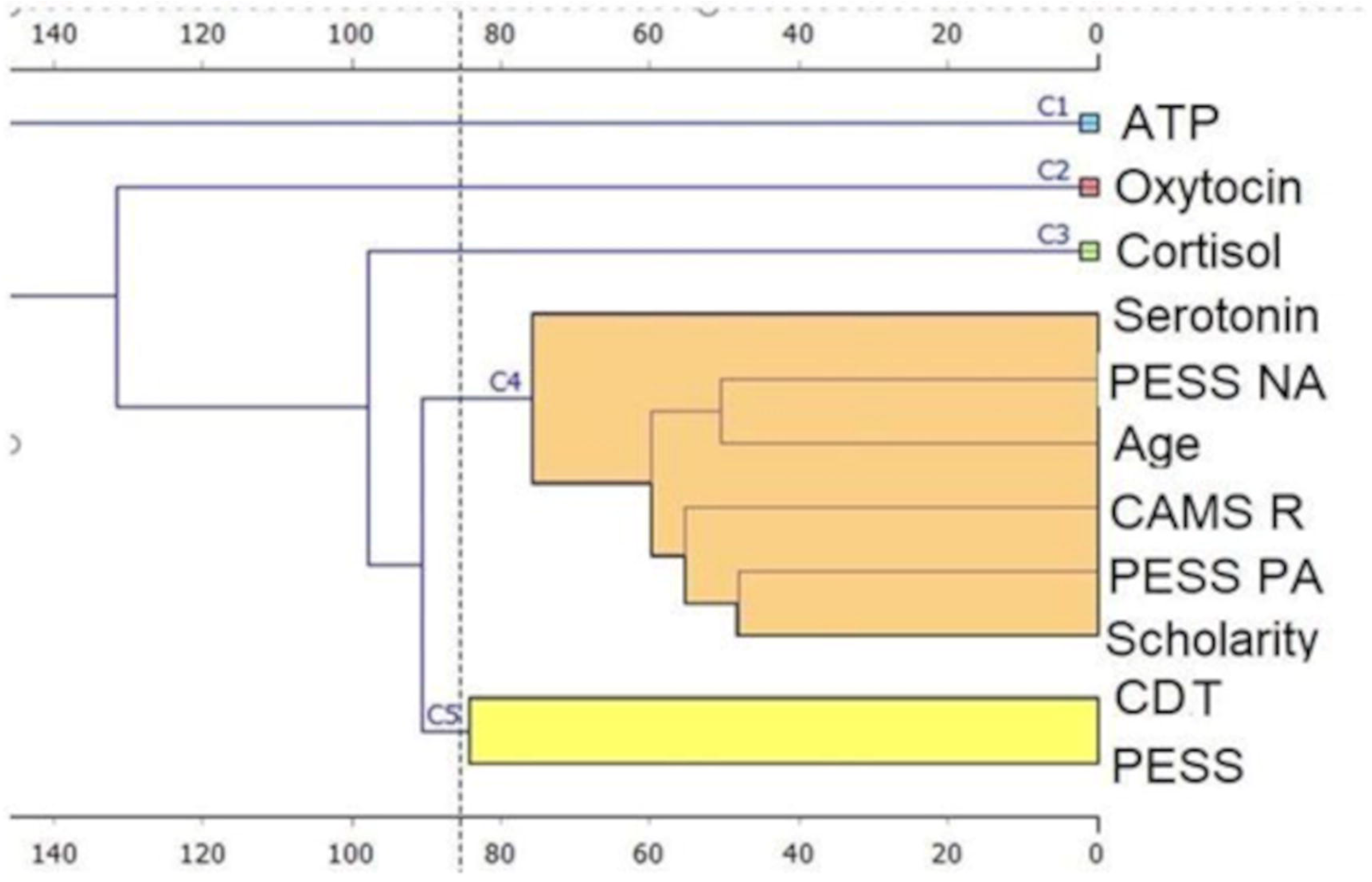
– Dendogram obtained for hierarchical cluster analysis by columns considering a single linkage algorithm and using the Manhattan distance on variables measured in CG and EG. CDT and PESS seem to have a closer relation (cluster C5) while another cluster appears when serotonin moderates the effect the association between clusters formed by the association of PESS NA and age (NA&age) and the association between CAMS-R with PESS PA and level of education ((PA&scholarity) & CAMS-R) (cluster C4). Cortisol changes showed to be associated to the clusters associated before (cluster C3), followed by oxytocin and ATP levels variations (clusters C2 and C1). CAMS-R, Cognitive and Affective Mindfulness measured by the Cognitive and Affective Mindfulness Scale-Revised, CDT, Cognitive functioning assessed by the Clock-Drawing Test, NA and PA, Affectivity assessed by the Positive and Negative Affect Schedule (PANAS), PESS, Emotional States assessed by the Perception of Emotional States Scale

## Discussion

The present study is a control compared, pre-post study of a time-limited intervention, aiming to identify any associations between biochemical/psychological changes upon clowning performances versus hosting conditions, in patients undergoing oncological treatment in ambulatory hospital rooms.

Findings in previous reported clown interventions with pediatric oncology patients undergoing chemotherapy, revealed a significant decrease of saliva cortisol levels, and improvement of psychological stress and fatigue in pre-post evaluation at four selected time points (48, 49). Although differences found were not statistically significant at all time points, the sample size was much reduced (n=6/16) and possible biases (e.g., sex) disregarded. Here, no significant reduced salivary cortisol is observed, but PdO intervention session is shorter, although with larger sample (n=64) and sex bias is valued. Other studies showed lower cortisol levels after clowns’ session with pediatric patients in acute conditions (e.g., 50), with clowns visits to two independent groups (n=18 patients each), for 90 minutes. In contrast, a pilot study conducted at a pediatric emergency department (51), the clowns reduced distress from venipuncture in 29 children (24 controls), which is identical to our groups’ size, but no effect on cortisol levels was observed. Nevertheless, independently of the PdO intervention, a significant decrease in cortisol levels (p=0.045) is observed, possibly linked with stress reduction in both groups that can be related with hosting conditions, since cortisol reduction was associated with relaxing environments (52).

Contrary to the present results, effective increase of children’s salivary oxytocin and lower anxiety ratings pre-post evaluated difference attributed to HCs intervention (5-120 minutes) was observed before (53), but not concerning emotional dimensions (n=17 (intervention), n=14 (controls)). Oxytocin was known to mediate positive affect and emotions, and sociability, providing chronic stress-deadening effects (54). Taylor (55) suggested that oxytocin system was involved in social affiliation through stress regulation, having an important biological role in stress-protective effects of positive social interactions (56). We undertake that PdO intervention is not influencing oxytocin-related functions in order to induce physiological changes, namely in social cognition and related behaviors for the groups in study. Moreover, oxytocin seems to correlate negatively to PA and CAMS-R in CG (Table 3), suggesting an involvement between these variables under hosting conditions only.

Regarding serotonin levels, no statistically significant differences are observed. Salivary serotonin was negatively linked with happiness in socially-positive settings (57), with individual’s current mood from collegiate runners before and after physical activity (58), which is in agreement with the trend of the results herein obtained (see also Figures 2 and S2). Furthermore, the diurnal rhythm of salivary serotonin concentration proved to differ between healthy individuals, who tended to have lower morning levels, and patients suffering from depression (59). The current data align with literature, and a link is found between serotonin variations and psychological changes, independently of PdO intervention, suggesting that serotonin may play a role in moderating the relationship between (PA&scholarity) and CAMS-R with (NA&age) (Figure 2). In CG, serotonin seems to have a role in psychological variations (more in PA and NA than CDT, PESS and CAMS-R) and linked to ATP levels (Figure S2, Table 3); in EG, serotonin is also somehow differently associated to these variables (Figure S2).

Salivary ATP levels do not show statistical significance in pre-post evaluation, seemingly, the HCs intervention stimulus is not sufficient to induce a mitochondrial bioenergetics alteration like ATP synthesis modification reflected in saliva. Still, ATP levels appear to be associated with psychological profiles and serotonin (Figure S2 and Table 3).

Biomarkers pre-post analyzed are located further away in clusters diagram of the patients evaluated, except for serotonin (Figure 2). Also, they organize not similarly in each one of the groups, due to ATP becomes nearly associated to other variables as serotonin (Figure S2). Additionally, higher ATP levels is significantly correlated with higher serotonin in CG, and with higher cortisol in EG in pre-post evaluation differences (Table 3). This suggests that biomarkers may be interlinked in the responses triggered, as a way to improve well-being conditions, involved as players in the HPA and SAM activities, and serotoninergic transmission. Besides, cortisol and oxytocin differences seem to be independent from the PdO intervention in their role of modulating changes of psychological status, serotonin and ATP levels (Figure S2).

The psychological status herein explored reveal improvements due to HCs performances, namely in negative affect (NA), positive emotional states (PESS) and mindfulness qualities (CAMS-R) of patients (Table 2). The intervention has a beneficial influence on NA in EG, compared to CG (p=0.032), meaning that the negative humor state of EG’s patients significantly decreases (e.g., distress, anxiety, fear, guilt). These results reinforce the conclusion of Casellas-Grau et al. (10), using clowning performances of 15 minutes with 99 adult cancer patients, helped to reduce the level of negative psychological symptoms (discouragement, anxiety, sadness, anger, worries, deception or fear) and fatigue feelings, also ameliorating the positive symptoms (feeling like laughing, happiness). Our results showed a tendency to be activated, enough to produce changes only in post-test difference between EG and CG (p=0.036). The CAMS-R values also increase from pre-to post-test in EG (p=0.037), suggesting the occurrence of a trend for improvement in attention, tolerance and acceptance of the present moment after intervention. The positive emotions (PESS) changes from pre-to post-intervention in EG, reflecting a better emotional functioning of these patients (p<0.001). The cognitive functioning, evaluated by CDT, is not significant in pre-post evaluation (p=0.442), indicating that the HCs intervention has no statistically significant impact on the learning effect.

Former research based on HCs visits during chemotherapy focused mainly in children, reporting higher levels of calm and happiness, less fatigue, pain, and distress, when compared to control group (n=41 both groups) (15). Moreover, these positive effects were higher in patients receiving HCs during two sessions, indicating that their intervention may extend for a longer effect on the well-being. In a different clinical context with children undergoing surgery, whom interacted with HCs during 15 minutes (n=35; n=35 controls), reported significantly less worries and showed improvement in emotional states, with decreased stress, suggesting that children are more predisposed to HCs interventions effects (60). A meta-analysis focused on the attempt to achieve a deeper psychological and shadings of the understanding of HCs encounters with ailing children suggested that the clowns meeting created the magical safe area, when children played and tested their possibilities (61). Children had the opportunity to recognize themselves in the state they were in at that exact moment and no longer focusing on their illness or limitations (1, 61, 62). Nevertheless, clowning intervention gained popularity in clinical care of adults and elderly (18). One of the first studies showed that HCs effectively led to lung function improvements of patients with chronic obstructive pulmonary disease, compared to controls (63). Others showed that HCs elicited amusement, feelings of transcendence, translated by feelings of being uplifted and surpassing the ordinary (64). Also, the clown effect of 1-day humor session followed by 9-12 humor-therapy sessions did not significantly reduce depression but significantly reduced agitation on a large group of elderly nursing home residents (65).

Our results are concordant with the above studies, meaning that HCs have a positive influence on psychological status of adult patients. The EG patients report high satisfaction with the HCs intervention (Supplements). Regardless, a follow-up intervention(s), an evaluation of a probable overcome in physical symptoms (e.g., fatigue) or improvement in results of disease treatment, which could reveal more details concerning the well-being of patients in analysis, were not performed.

The lack of significant differences in EG positive affect can be due to the fact that the evaluation subscale used terms referring states related to a high level of excitement, and which could be interpreted by patients as such and then be underappreciated, bearing in mind their illness context (e.g., feeling enthusiasm, inspiration). This explanation is consistent with the improvements in positive emotional states evaluated by PESS, since it captures emotional functioning easier to change with performances, such as feeling calmer, joyful, relaxed and communicative. Theoretically, the affective component of subjective well-being (PA and NA) requires high levels of positive affect and, conversely, lower levels of negative affect (5). Considering these dynamics, the empirical data suggest that PdO brief performances impacts on oncological patients’ well-being, mainly via decreasing negative psychological symptoms (they are living a condition of highly distress), more than improving positive affect, although the emotional states more aroused by what is happening in the moment also improved. In fact, NA is highly positively correlated to PESS in EG pre-post evaluation (Table 3), suggesting that there may be a mutual change of both parameters. Moreover, PA and CAMS-R share a link (Table 3).

Furthermore, there are no significant differences in all psychological aspects for CG, although variations in CDT, NA, PESS and CAMS-R all had a beneficial tendency (table 1). Accordingly, a normal oncological treatment session is not marked by stressful triggers for the patients, due to the supportive room environment, and it could reflect the quality of the interaction between the evaluation team and the patients since it was marked by kindness, active listening, availability, further reinforcing the good treatment atmosphere. This result is reinforced by the observed global decrease in cortisol, mentioned previously, in pre-post evaluation.

Although statistically significant differences were verified between groups in the pre-test evaluation of psychological features of PESS (p=0.014) and NA (p=0.042), it is assumed that this fact occurred by chance in the distribution into small size groups.

The emotional state and perception may be influenced by age and scholar level. The variation in PA is modulated by the education level (PA&scholarity), which is associated with CAMS-R (Figure 2). This is reinforced in EG as seen in Table 3 and Figure S2). It may strengthen the aspect of PA variation in EG, although not significantly different to CG per se (Table 2). The education level factor here may be influencing this relationship. Additionally, this cluster is attached to NA&age (Figure 2), suggesting that the observed significant decreasing of negative mood states (NA) of patients is moderating the effect of the improvement in attention, tolerance and acceptance in and of the present moment (CAMS-R) and positive affect (PA) relationship. Notably, as already noted that salivary serotonin is linked inversely with happiness and current mood (57, 58), it may play a role in moderating the relationship between those clusters (Figure 2), suggesting that the observed significant improvements in these psychological features is associated with this biomarker drifts, independently of the HCs intervention. Figure S2 shows that serotonin has a role in psychological profiles, closer to PA and NA, only emerging in CG. Serotonin and ATP levels’ differences are also closely related to CDT and CAMS-R differences in both groups (Figure S2), but in EG, a trade between scholarity and PESS occurs, evidencing a turn to a modulating role of the emotional feelings to the other psychological features changes and serotonin and ATP changes of EG.

The attempt to comprehensively examine the effectiveness of the HCs (PdO) intervention for enhancing emotional and behavioral processes using biomarkers, may be further ameliorated. Some studies might indicate clues for these approaches (49–51). Most likely it would take a higher number of sessions and/or longer HCs sessions for changes to occur, or even increasing the sample size, by perhaps doing collection of more than one saliva samples in pre– and post-interventions in order to obtain a more differentiating effect between groups. However, the particular circumstances of the patients in study may also contribute to the results obtained.

Although presenting some limitations, such as the fact that the population in study included adult women with a wide age range being treated for different types of cancer, the novelty of analyzing well-being biomarkers is undeniable. Certainly, additional research that would include males, narrower age groups with higher number of subjects and stratified according to the subtypes of cancer is an open field to explore. The sample size did not allow the subdivision of data to explore deeper additional factors, like age and educational level’s effects.

Despite the recognition of HCs as supporting agents in health care settings, this study does not show changes in the biomarkers studied, possibly because of the short duration of its intervention or related to the nature of the intervention that affect immediate emotional states, but do not influence the cognitive functions. It is remarkable to notice that the good hosting conditions of Hospital are reducing patients’ stress, which is essential for the treatment success and well-being of the patients in an ambulatory chemotherapy hospital setting.

In conclusion, psychological functioning evidences observed show that short-term HCs interventions effects during ambulatory chemotherapy seem to contribute to increasing the well-being of adult cancer patients, reinforcing the importance of this kind of interventions in ambulatory treatment conditions environment. Future randomized research studies with higher control levels and larger samples are needed, to both ascertain psychological benefits and salivary biomarkers variations, and consolidate the relevant variables.

Additionally, although preliminary, due the nature of the study (pilot), it is important to stress out the novelty of this work, in which a set of well-being biomarkers was implemented, in correlation with psychological evaluation including cognitive function, besides emotional states, and all evaluated by cluster analysis. This experimental approach can be used in any study examining the effect of any type of intervention possibly related to well-being in mental health studies, HCs or any other artistic/social interventions, sports, disease settings or work environments.

## Data Availability

All data produced in the present study are available upon reasonable request to the authors

## Acknowledgements

The authors especially grateful the remarkable dedication of health professionals from the Portuguese Institute of Oncology of Coimbra (IPO) (Instituto Português de Oncologia de Coimbra Francisco Gentil, E.P.E. (IPOCFG, E.P.E.)), with special thanks to Dr. Margarida Ornelas (President of the Administrative Council), Dr. Ana Pais (Clinical Director), and to Dr. Gabriela Sousa (Director of Medical Oncology Service). The authors are especially grateful to Vanessa Costa (former MSc student at LBioMiT), for all her efforts and relevant contribution to the development of this study. The authors also acknowledge the clowns from “Palhaços D’Opital” for their collaboration in the performance for the intervention to the experimental group (EG). The authors truly thank all women for their participation in this study, which allowed the accomplishment of the project aims.The authors are thankful to Paulo Matafome for the fruitful discussions concerning ELISA and to Professor Teresa Gonçalves and Chantal Fernandes for the availability of using the lyophilizer (serotonin evaluation).

This work was partially financed by European Social Fund, as part of the Portugal 2020 Partnership Agreement, within the scope of Portugal Social Innovation initiative (POISE-03-4639-FSE-000427). Additional support was granted by the European Regional Development Fund (ERDF), through the Centro 2020 Regional Operational Programme under project CENTRO-01-0145-FEDER-000012-N2323 and CENTRO-07-ST24-FEDER-002002/6/ and through the COMPETE 2020 – Operational Programme for Competitiveness and Internationalization and portuguese national funds via FCT – Fundação para a Ciência e a Tecnologia, under project[s] POCI-01-0145-FEDER-007440, and UIDB/04539/2020, UIDP/04539/2020 and LA/P/0058/2020.

