## Supplemental data for "Well-being biomarkers and psychological functioning of adult patients during chemotherapy treatment: the effects of hospital clowns and hosting conditions"

Supplemental Information

**Methods and Materials**

**Study design**

Pre- and post-test procedures included the saliva collection followed by the psychological assessment. The schedule was as follows: 09.00 a.m. welcome, study presentation and informed consent signature; 09.30-10.40 a.m. saliva collection time 1; 09.40-10.40 a.m. psychological evaluation time 1; 10.40 a.m. HCs intervention for EG and without for CG; 11.00-12.10 a.m. saliva collection time 2; 11.15-12.15 a.m. psychological evaluation time 2. The planned schedule time has taken into account the chronobiology of cortisol as reference, since serum cortisol levels are highest between 7 a.m. and 8 a.m. and during the day, the levels of cortisol drop significantly and, in the evening, only about 10% of the morning cortisol remains in the body (1).

**Participants, Recruitment, and Procedures**

This study was carried out in the Portuguese Institute of Oncology of Coimbra (IPO) and it was approved by the respective Ethics’ Committee (#03PI/2020 IPO), after authorization given by the Directors of IPO, Clinical Board and Service (ambulatory hospital) in which the saliva collection and psychological evaluation were conducted.

The recruitment of participants was carried out by a nursing professional, according to the appointments of patients whom would be receiving the usual chemotherapy treatment in ambulatory hospital setting, on the days defined for data and samples collection. The exclusion criteria included patients with: head and neck cancer; having their first or last chemotherapy session; receiving chemotherapy orally or via lumbar puncture; and presenting with any cognitive or language impairment that would prevent their understanding of the study and assessment or that could predictably interfere the levels of biomarkers evaluated.

Participants were told that they would participate in a study to better understand the variations in the relationships between saliva biomarkers’ properties and psychological emotional and attentional states across morning time and those who agree to participate were provided with written informed consent at arrival to ambulatory hospital.

**Samples collection**

Before saliva collection, all participants were instructed to avoid foods with high sugar or acidity or high caffeine content, immediately before sample collection, as it could compromise the test by decreasing the pH of the saliva or by increasing bacterial growth. They were also requested to rinse their mouths with water to remove any food residue and to wait at least 10 minutes after rinsing, prior to collection, in order to avoid sample dilution.

Saliva collections were performed by the participants themselves, following instructions and using Saliva Collection Aid Salimetrics (5016.02-SAL). The samples were collected according to the schedule as described above and stored at 4°C. The two saliva samples harvests were separated by about 90 minutes (time lapse related to the data collection for psychological evaluation and the length of time with the social intervention by HCs). Then, samples were transported to the laboratory, in order to be centrifuged and stored at -80°C for further analysis.

**Determination of well-being and bioenergy biomarkers’ levels**

The evaluation of well-being biomarkers, oxytocin, cortisol and serotonin, was performed by using ELISA technique (Enzyme-Linked Immunosorbent Assay), because it allows the quantification of low levels of various analytes, in different types of biological samples, with high sensitivity. The measurement of the concentration of each biomarker was performed using specific kits for each analyte.

In the case of oxytocin, extraction was required prior to ELISA analysis, using SPE columns, acetonitrile and trifluoroacetic acid (TFA), according to manufacturers’ instructions. A 200 mg C18 Sep-Pak column was equilibrated with 1ml of acetonitrile, followed by 10 mL of 0.1% TFA-H2O. Then, the saliva supernatant was applied to the column and washed with 10 mL of 0.1% TFA-H2O. Finally, the sample was slowly eluted (gravity-fed) by applying 3mL of 95% acetonitrile/5% of 0.1% TFA-H2O solution. The eluate was collected into a plastic tube and evaporated to dryness under nitrogen gas. Oxytocin levels in extracted saliva were measured using the Enzo Life Sciences kit (ADI-900-153A). In the first phase of this method, the endogenous oxytocin competes with oxytocin linked to alkaline phosphatase for the oxytocin antibody binding sites. After overnight incubation at 4°C, the excess of reagents was washed away and the bounded oxytocin phosphatase was incubated with the substrate. After 1h, this enzyme reaction that generates a yellow color and it was stopped with a Tris-sodium solution. All the samples were analyzed in duplicate and the optical density was red in a spectrophotometer at λ=405nm (Spectra Max plus 384, Molecular Devices). Finally, the saliva oxytocin content (pg/mL) was determined by plotting the optical density of each sample against the standard curve (2). The sensitivity of quantification was 15pg/mL (range 15.6 - 1,000pg/mL), the intra-assay precision was 12.6 (%CV) for 39.9pg/mL control, 10.2 (%CV) for 121.4pg/mL control, 13.3 (%CV) for 363.7pg/mL control, and the inter-assay precision was 20.9 (%CV) for 47.0pg/mL control, 16.5 (%CV) for 145.1pg/mL control, 11.8 (%CV) for 397.2pg/mL control.

Concerning cortisol evaluation, the samples were thawed and centrifuged for immediate immunoassay processing with ADI-900-071 (Enzo Life Sciences) kit. This assay uses a monoclonal antibody to competitively binding cortisol, covalently attached to an alkaline phosphatase molecule. After incubation at room temperature, the excess of reagents was washed away and substrate was added. After a short incubation time, the enzyme reaction was stopped with a Tris-sodium solution and the optical density was red at λ=405nm. A standard curve was used to estimate the cortisol content (pg/mL) (2). The sensitivity of quantification was 56.72pg/mL (range 156 - 10,000pg/mL), the intra-assay precision was 10.5 (%CV) for 333pg/mL control, 6.6 (%CV) for 1,088pg/mL control, 7.3 (%CV) for 3,155pg/mL control, and the inter-assay precision was 13.4 (%CV) for 451pg/mL control, 7.8 (%CV) for 969pg/mL control, 8.6 (%CV) for 3,052pg/mL control.

In the serotonin determination, samples were concentrated by lyophilization. The samples were frozen in liquid nitrogen and then processed in a freeze dryer. The serotonin ELISA analysis was performed using a colorimetric competitive enzyme immunoassay (ADI-900-175 (Enzo Life Sciences)) kit, according to manufacturers’ instructions. In brief, the samples were applied to a goat anti-rabbit polyclonal antibody coated plate, followed by adding solutions of a serotonin-alkaline phosphatase conjugate and a rabbit polyclonal antibody to serotonin. After the antibody competitively binding, plate was washed for remaining with only bound serotonin, which reacted to substrate added for alkaline phosphatase catalysis. A Tris-sodium solution was added to stop the reaction. The optical density was red at λ=405nm and the serotonin content (ng/mL) was determined by plotting the optical density of each sample against the standard curve (2). The sensitivity of quantification was 0.293ng/ml (range 0.49 - 500ng/ml), the intra-assay precision was 11.0 (%CV) for 13.5ng/mL control, 5.8 (%CV) for 53.8ng/mL control, 4.2 (%CV) for 346.1ng/mL control, and the inter-assay precision was 12.7 (%CV) for 13.2ng/mL control, 18.4 (%CV) for 53.9ng/mL control, 16.2 (%CV) for 358.0ng/mL control.

The assessment of the saliva ATP levels was performed using a bioluminescent assay (ATP Bioluminescence Assay Kit HS II, Roche Diagnostics), in a Luminometer Berthold FB12. This technique uses the ATP dependence on oxidation catalyzed by luciferase and allows the measurement of extremely low concentrations of ATP (up to 10^-15^ mol) in the saliva samples (3). The protein concentration was assayed by the Bradford method (4), at λ=595nm (Spectra Max plus 384, Molecular Devices) and ATP levels were expressed normalized to protein concentration for the sake of results’ accuracy.

A portion of collected saliva samples (n= 1 – 6 in CG and n= 5 – 9 in EG, depending on the biomarker) was excluded for the biomarkers’ levels assays because they either had insufficient saliva or because their baseline biomarkers’ levels were too low to be measured.

**Psychological evaluation**

*Affectivity* was assessed by the Positive and Negative Affect Schedule (PANAS) developed by Watson et al. (1988) (5) and adapted to Portuguese by Galinha et al. (2014) (6). The PANAS has two independent subscales with five items each. Responses are in a Likert scale and can vary from 1 (“Nothing or very slightly”) to 5 (“Extremely”). The PANAS can measure affective and mood states in the moment. Higher values in positive affect (PA) and lower values in negative affect (NA) correspond, respectively, to a better affectivity. The Portuguese adaptation showed good psychometric properties, internal consistency (Cronbach’s alpha =0.86 for PA) and 0.89 for NA) and structural validity through Confirmatory Factor Analysis.

*Emotional States* were assessed by the Perception of Emotional States Scale (PESS), adapted from Tuckman (2012) (7) based on semantic differential scales and consisting of 8 pairs of opposite adjectives separated by 7 positions (e.g., calm/nervous; happy/sad). These pairs were selected because they represent emotional states, different from the affective states assessed by PANAS. Lower ratings correspond to a better emotional state.

*Cognitive and Affective Mindfulness* was measured by the Cognitive and Affective Mindfulness Scale-Revised (CAMS-R) originally developed by Feldman et al. (2007) (8). The scale was adapted by Teixeira et al. (2017) (9), having 9 items rated to a 4-point Likert scale, ranging from 1 (Not at all) to 4 (A lot) and measures mindfulness qualities of attention, tolerance and acceptance of the present moment, with higher values corresponding to better results. Original and adaptation studies shown a stable “one-factor” structure and an adequate internal consistency (Cronbach’s alpha =0.76). For the present study, only 6 items were considered, because of the need to make the psychological assessment as brief as possible so that a minimum interference with ambulatory sessions functioning (e.g., lunch time) would occur and respecting the patient’s availability could be ensured.

*Cognitive functioning* was assessed by the Clock-Drawing Test (CDT, with higher values corresponding to better cognitive functioning). The CDT measures visuospatial, visuoconstructive and visuomotor capabilities. This test also evaluates other cognitive functions, including language/understanding, memory, attention and executive functions (planning, organization, simultaneous processing, self-monitoring). It is commonly used as a brief cognitive screening test. The Portuguese version of the 18-point quantitative scoring system was used (10, 11).

Satisfaction with HCs’ intervention was evaluated through two single items: (1) “Do you think this type of interventions in the hospital is important?”, answered on a 4-point Likert scale (ranging between "Very important" and "Nothing important"); and (2) "How satisfied are you with the intervention of the HCs that you just saw?” answered in a 5-point Likert scale (ranging between "Very Satisfied" and "Very Unsatisfied").

**Statistical analysis**

Qualitative data were described by its absolute and relative frequency while mean ± standard deviation and median [Quartile 1; Quartile3] were presented for quantitative data.

A factorial repeated measures ANOVA was applied in order to evaluate either the effect of evaluation moments and group interaction, and the main effect of the repeated measure (although some variables have presented normality deviations on their residuals, that fact did not have a significant impact on lowering power on those analysis). Moreover, bivariate comparisons between evaluations in each group were performed either using a Student’s t-test for paired samples or a Wilcoxon test, according to sample distribution, and between groups for each time point using an independent samples t-test or a Mann-Whitney U-test using the same criteria. Pearson and Spearman rank order correlations between pre-test and post-test difference in biomarkers and psychological characteristics were also obtained for each group, and they were evaluated according to each data distribution. Furthermore, the differences of the changes in time of the variables were evaluated in function of the age and educational level of the patients between the two groups using a repeated measures ANOVA considering those as factors in a full factorial model.

These analyses were performed in IBM SPSS, version 25, and were analyzed at a 5% significance level (at a 10% significance level concerning Pearson and Spearman rank order correlations).

In order to assess association between biomarkers and psychological characteristics, cluster analyses using a Manhattan distance with single linkage were performed in Orange version 3.28.0 and the dendograms obtained were presented.

**Results**

**Table S1** - Sociodemographic data, including age, educational level, marital status and profession

| Patients | Control Group (CG) | Experimental Group (EG) |
| --- | --- | --- |
| n | 28 | 36 |
| Age  Mean (SD)  Min - Max  p=0.655 (mean age between groups) | 56.7 (9.9)  36 - 77 | 58.1 (13.5)  32 - 90 |
| Educational level (n (%))  No studies  Up to 4^th^ year of school  6^th^ year of school  9^th^ year of school  12^th^ year of school  Higher school | 0 (0%)  9 (32.1%)  6 (21.4%)  5 (17.9%)  4 (14.3%)  4 (14.3%) | 1 (2.8%)  10 (27.8%)  6 (16.7%)  5 (13.9%)  5 (13.9%)  9 (25.0%) |
| Marital status  Single  Married/partnered  Widowed  Divorced/separated  NA | 1 (3.6%)  17 (60.7%)  3 (10.7%)  7 (25.0%)  0 | 1 (2.8%)  23 (63,9%)  9 (25.0%)  2 (5.6%)  1 (2.8%) |
| Profession (working status)  Employed  Pensioner  Unemployed | 22 (78.6%)  4 (14.3%)  2 (7.1%) | 28 (77.8%)  7 (19.4%)  1 (2.8%) |

NA – not answered.

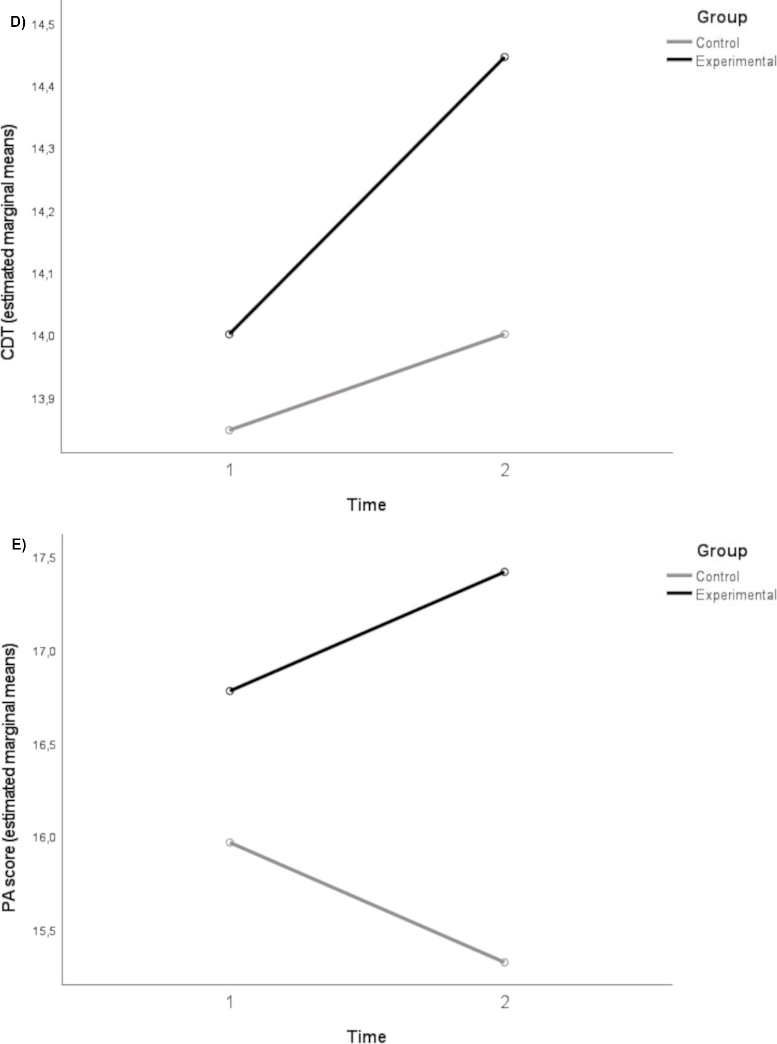

**Figure S1** – Psychological functioning variation between pre- and post-test evaluations for both Control (CG) and Experimental groups (EG). D) CDT (no statistically significant variation found in CG – p = 1.000, and EG - p=0.442 (see Table 2); E) PA (no statistically significant variation found in CG – p = 0.211, and EG - p=0.277 (see Table 2)). CDT, Cognitive functioning assessed by the Clock-Drawing Test, PA, *Affectivity* assessed by the Positive and Negative Affect Schedule (PANAS).

When considering the evaluation of the pre- and post-test variations in the levels of biomarkers and psychological features, according to individuals’ age and education levels, there was no interaction detected in age subgroups or according to scholar degrees.

Separate analyses suggest that psychological features associated with serotonin and ATP levels differences of the CG might be modulated by educational level (Figure S2A). In EG, a trade between scholar level and global PESS differences occurs, but serotonin and ATP levels differences keep closely related to PA and NA differences, CDT and CAMS-R (Figure S2B). Oxytocin levels, age and cortisol levels differences seem to be independent from the intervention in their role modulating previously observed association patterns; however, oxytocin is closer to the other features than cortisol or age.

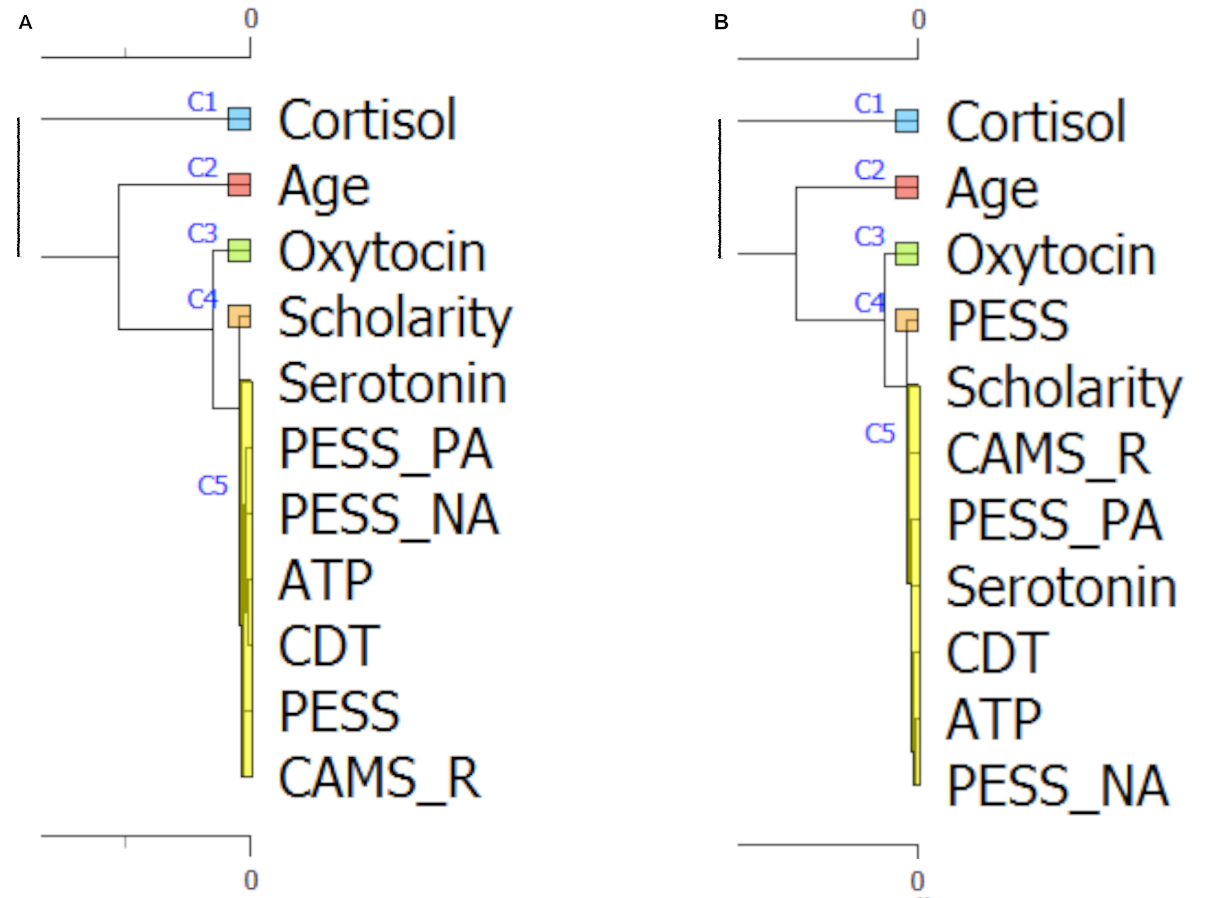

**Figure S2** - Hierarchical cluster analysis by columns on variables measured in the CG (A) and in the EG (B). As observed, the organization in each one of the groups is not similar; in EG ATP levels seem to be associated with psychological profile while in the CG serotonin has also a role in that, although more specifically just in PA and NA.

CAMS-R, Cognitive and Affective Mindfulness measured by the Cognitive and Affective Mindfulness Scale-Revised, CDT, Cognitive functioning assessed by the Clock-Drawing Test, NA and PA, Affectivity assessed by the Positive and Negative Affect Schedule (PANAS), PESS, Emotional States assessed by the Perception of Emotional States Scale.

**Satisfaction with HCs intervention**

Concerning patient’s satisfaction with HCs intervention (EG), the obtained results pointed mainly to a very important performance and to high satisfaction with this kind of art events in the hospital setting. Patient’s answers to the question “Do you think this type of interventions in the hospital is important?” were rated as “very important” (75%, n=27) and “important” (25%, n=9). Moreover, responses to the question “How satisfied are you with the intervention of the HCs that you just saw?” were scored as “very satisfied” (51.4%, n=18), “satisfied” (42.9%, n=15) and only 2 patients selected the “indifferent” option (5.7%). Reinforcing the appreciation of this type of intervention by cancer patients, qualitative data collected by the evaluation team concerning self-assessment of the participants' reactions to the HCs’ intervention, showed that the four most mentioned reactions were “smiles”, “I joined in the games”, “I talked to the Clowns”, “I contributed to the good atmosphere in the room” and “I laughed”. These indicators suggest that oncological patients’ satisfaction with this type of events is high and a residual number of patients is indifferent.
